# SARS-CoV-2 specific memory B cells frequency in recovered patient remains stable while antibodies decay over time

**DOI:** 10.1101/2020.08.23.20179796

**Authors:** A. Vaisman-Mentesh, Y. Dror, R. Tur-Kaspa, D. Markovitch, Tatiana Kournos, D. Dicker, Y. Wine

**Affiliations:** The Shmunis School of Biomedicine and Cancer Research, George S. Wise Faculty of Life Sciences, Tel Aviv University, Tel Aviv 69978, Israel.; Department of Medicine D and the Liver Institute, Rabin Medical Center, Beilinson Hospital, Molecular Hepatology Research Laboratory, Felsenstein Medical Research Center, Sackler School of Medicine, Tel Aviv University, 39 Jabotinsky Street, Petah-Tikva 49100, Israel; Internal Medicine D, Hasharon Hospital-Rabin Medical Center, Petach Tikva, Israel. Sackler School of Medicine, Tel Aviv University, 6997801 Israel

**Keywords:** COVID-19, SARS-CoV-2, B cell response, Antibody, serology, antibody repertoire, RBD, humoral response, serological memory

## Abstract

The breadth of the humoral immune response following SARS-CoV-2 infection was indicated to be important for recovery from COVID-19. Recent studies have provided valuable insights regarding the dynamics of the antibody response in symptomatic COVID-19 patients. However, the information regarding the dynamics of the serological and cellular memory in COVID-19 recovered patients in scarce. It is imperative to determine the persistence of humoral memory in COVID-19 recovered patients as it will help to evaluate the susceptibility of recovered patients to re-infection. Here, we describe the dynamics of both the SARS-CoV-2 specific serological and B cell response in COVID-19 recovered patients. We found that symptomatic SARS-CoV-2 patients mount a robust antibody response following infection however, the serological memory decays in recovered patients over the period of 6 months. On the other hand, the B cell response as observed in the SARS-CoV-2 specific memory B cell compartment, was found to be stable over time. Moreover, the frequency of SARS-CoV-2 specific B cell plasmablasts was found to be associated with the SARS-CoV-2 specific antibody levels. These data, suggests that the differentiation of short-lived plasmablasts to become long-lived plasma cells is impaired and the main contributor of antibody production are the short-lived plasmablasts.

Overall, our data provides insights regarding the humoral memory persistence in recovered COVID-19 patients. Notwithstanding the insights from this study, it is still to be determined if the persistence of SARS-CoV-2 memory B cells can be considered as a correlate of protection in the absence of serological memory.

## Background

The first patients detected positive to coronavirus disease 2019 (COVID-19) were identified in Wuhan, China^1^. These patients were found to be infected by severe acute respiratory syndrome coronavirus 2 (SARS-CoV-2) leading to the declaration of the World Health Organization (WHO) that COVID-19 is a worldwide pandemic^2^. Rapid response to the outbreak provided important information regarding the virus genome sequence and especially the Spike protein (S protein) and its sub-region, the receptor binding domain (RBD) which is responsible for the binding to the human host cell receptor, human Angiotensin-converting enzyme 2 (hACE2) and mediates virus entry^3^. The S protein is highly immunogenic with the RBD holding the potential to elicit neutralizing antibodies (nAbs)^4^. The S protein RBD, was shown to elicit antibodies starting early as 3 days following the onset of symptoms^5^ in which their levels increase along the progression of the disease^6–8^. Measuring RBD-specific serum antibody titers in samples collected from COVID-19 patients during active disease, suggests that in most cases, SARS-CoV-2 infection mounts a robust humoral response and generates antibodies specific to the S protein RBD^9^. Many serological studies are focused on developing diagnostic approaches for the detection of virus-specific antibodies by the identification of IgM and IgG antibodies as they are indicative of a productive progression of humoral immune response. Serological tests for specific SARS-CoV-2 antibodies detection in patient’s blood are important to trace contacts and for serological surveillance. Still the question remains if serological memory persist over time and recent studies suggest that the antibodies mounted against SARS-CoV-2 do not persist over time and decay several weeks following onset of symptoms^10^. At the cellular level, the T cell population was investigated as it was suggested that these cells may contribute to recovery^11^. T cell reactivity to SARS-CoV-2 epitopes was detected in non-exposed individuals suggesting a cross-reactive T cell recognition between circulating “common cold” coronaviruses and SARS-CoV-2^12^. The latter report further strengthens the idea that memory wanes over time as the population does not develop immunity to common cold. On the other hand, SARS-CoV-2 specific T cells present in the general population may impact susceptibility and pathogenesis of SARS-CoV-2 infection^13,14^.

Recent studies report on the isolation of RBD-specific B cells aiming at elucidating the composition of neutralizing antibodies using single cell analysis^15,16^. However, the information regarding SARS-CoV-2 specific B cell dynamics is scarce and there is little information regarding the dynamics of antibodies and their association with the antigen-specific B cell compartment in COVID-19 recovered patients. This information is detrimental as it can help to evaluate the breadth of the humoral memory in COVID-19 recovered patients and to determine whither this will contribute for long-lasting serological and cellular memory. Adaptive memory is two-parted, comprising long-lived antibody-secreting plasma cells (LLPC) which contribute to the serological memory and long-lived memory B cells (mBC) that have the capacity to react quickly to a recurrent antigenic challenge (cellular memory)^17^. In this study we aim to understand the temporal dynamics of the serological and cellular memory in COVID-19 recovered patients.

To this end, we collected sera from 54 active COVID-19 patients and whole blood from 57 COVID-19 recovered patients and six follow up samples from the recovered cohort, along with their clinical data. Sera were tested to determine the levels of RBD-specific antibodies and their neutralization capacity. B cells isolated by FACS from COVID-19 recovered patients were used to determine the frequency of RBD-specific B cell subsets. We found that RBD-specific antibodies increase rapidly in symptomatic COVID-19 patients and further decline rapidly in recovered patients. We identified a strong correlation between the RBD-specific IgG levels and their neutralization capacity suggesting that RBD-specific antibody titer by itself may be used as a proxy to evaluate their neutralization capacity. Interestingly, we found that COVID-19 patients with metabolic disorder exhibit significant higher levels of RBD-specific antibodies.

Additional association was found between the levels of RBD-specific antibodies with ferritin and C-reactive protein (CRP) indicating that disease severity leads to a robust immune response. More importantly, we found that there is a correlation between the time following onset of symptoms and the RBD-specific antibody titers indicating that the recovered COVID-19 patients experience a decay in their serological memory. In contrast to this decay, RBD-specific memory B cells were found to be stable within the range of six months following onset of symptoms. Overall, our data obtained in this study reveals a discordance between serological and cellular memory in COVID-19 recovered patients which has important implications on reinfection susceptibility and vaccine design.

## Results and discussion

To study the acute phase antibody response to SARS-CoV-2 infection and the persistence of antibodies in recovered COVID-19 patients, RBD-specific IgG/A/M levels were measured in sera from symptomatic (n = 54), recovered (n = 57) COVID-19 patients and control sera (n = 26). Antibody measurements were carried out in duplicates and each experiment was repeated independently twice with high reproducibility (Figure S1). Antibody levels were calculated as the signal over mean signal obtained in the negative controls used in each microwell plate (Figure 1A). We observe that IgG/A/M levels in the symptomatic COVID-19 cohort, increases significantly 5 days following onset of symptoms (DFS). The antibody titers for all isotypes continues to increase to reach significantly higher levels compared to the control group approximately 15 DFS. IgG/A/M levels in recovered patients decreased significantly compared to the antibody levels in symptomatic patients (15-60 DFS) while still showing scientifically higher levels compared to the control cohort. IgA levels decreased rapidly in the recovered patients reaching the basal levels as observed in the control group.

**Figure 1.**
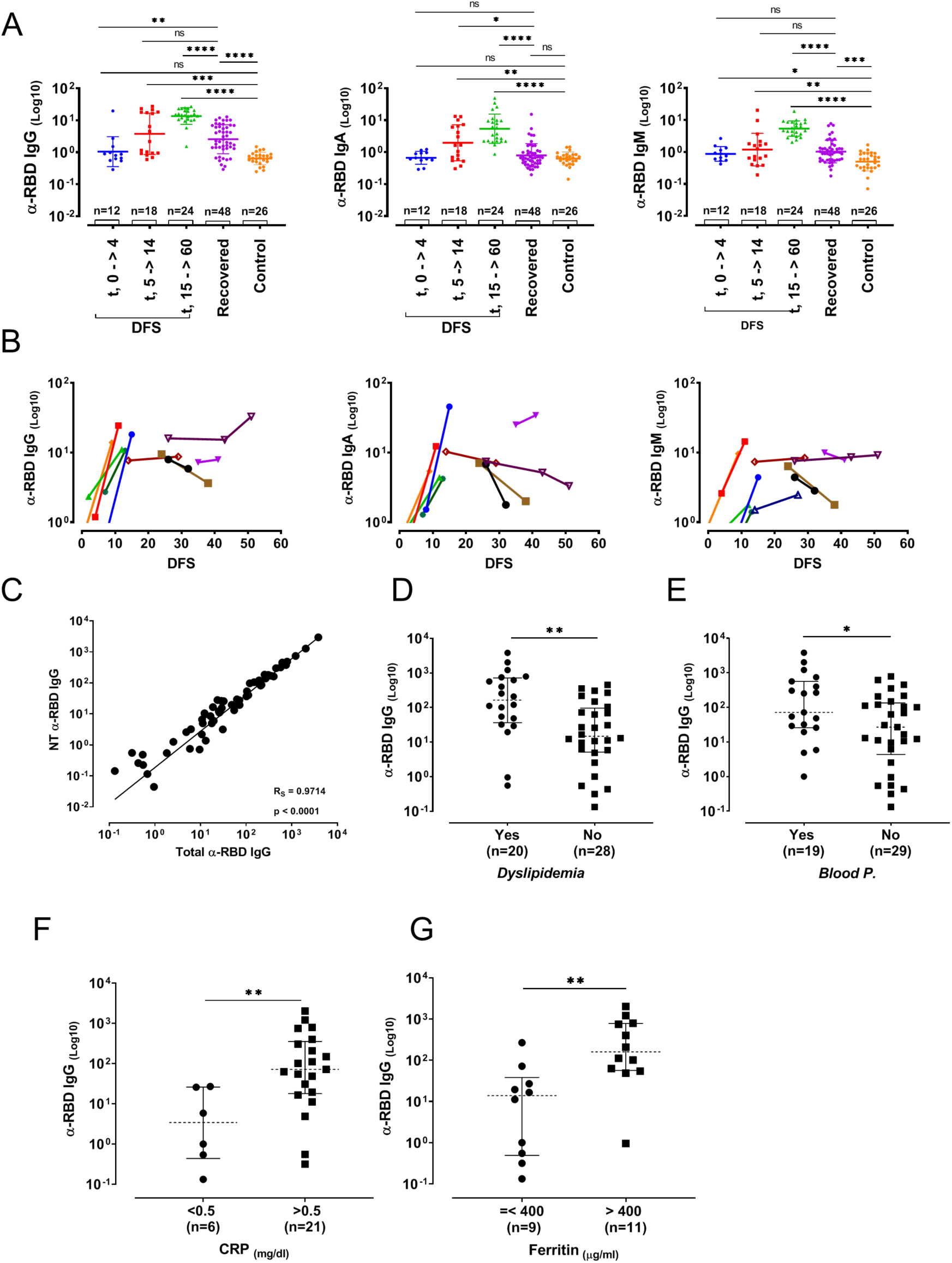
RBD-specific antibodies in symptomatic, recovered and control cohorts and their association with clinical parameters. (**A**) anti-RBD IgG/A/M antibody levels in symptomatic, recovered COVID-19 patients and control sera. Symptomatic COVID-19 patients were stratified into DFS groups (0–4, 5–14 and 15–60, the maximum value for DFS in symptomatic COVID-19 patients). Statistical significance was calculated using unpaired, nonparametric, two-tail P values, Mann Whitney Test. Geometric mean and geometric SD are shown with lines for each group. (**B**) RBD-specific IgG/A/M levels in follow up samples collected from a subset of symptomatic patients (n = 11). Sera samples were collected at two time point with one patient at three time points. X-values represent DFS. Each color designates a different patient. (**C**) Correlation between the levels of total RBD-specific antibodies and neutralizing antibodies in COVID-19 recovered patients sera using spearman’s rank correlation (n = 57). RBD-specific IgG levels in COVID-19 recovered patient sera, stratified by the metabolic syndrome factors, (**D**) dyslipidemia and (**E**) hypertension. RBD-specific IgG levels in COVID-19 recovered patient sera, stratified by COVID-19 risk factors (**F**) CRP and (**G**) ferritin. (ns p > 0.05, * p ≤ 0.05, ** p ≤ 0.01, *** p ≤ 0.01, **** p ≤ 0.0001).

Follow up sera were collected from 11 symptomatic patients to monitor the antibody dynamics over time within each patient (Figure 1B). We observed a rapid increase in RBD-specific antibody levels within 10 DFS for all isotypes which plateaued or started to decline, approximately 15 DFS. RBD-specific IgG titers in recovered COVID-19 patients, were measured also in dilution series and calculated as the ratio between area under curve (AUC) and the mean AUC of control sera that was applied to each microwell plate. Utilizing *in vitro* neutralization assay as described previously^18^ the nAb fraction out of the total RBD-specific antibodies was calculated. Each serum sample was applied to microwell plate w/ and w/o hACE2 as a competitive agent. Hence, the decrease of the signal is associated with the portion of nAbs that were depleted from the microwell due to blockade of epitopes on the RBD by hACE2 (Figure S2). We found a strong correlation between the total RBD-specific antibodies and the nAb suggesting that measuring RBD-specific antibody levels can be used as a proxy to evaluate the nAbs levels (Figure 1C).

Collected clinical parameters for a subset of the recovered patients were stratified by the clinical indications. It was previously demonstrated that dyslipidemia and high blood pressure are linked to metabolic syndrome^19^. We found that both parameters were associated with the levels of anti-RBD IgG (Figure 1D-E). C-reactive protein (CRP), hyperferritinemia and D-dimer were described as features of systemic inflammatory reactions in COVID-19 patients and were suggested to be used as an indication for disease severity^20–23^. Similar to the previous reports, we found that ferritin and CRP are correlated while ferritin and lymphocyte counts are inversely correlated (Figure S3). We examined the association between these COVID-19 severity risk factors and the levels of RBD-specific antibodies. Both serum ferritin and CRP were found to be associated with high levels of anti-RBD IgG (Figure 1F-G, Figure S4A-B) while elevated D-dimer was not found to be associated with increased levels of RBD-specific antibodies (Figure S4C). Age (n = 52, Figure S5A) and gender (Figure S5B) showed no, or weak correlation with RBD-specific IgG levels.

To evaluate the persistence of anti-RBD antibodies following COVID-19 recovery, the antibody levels were tested for their association with DFS. We found that the antibody levels were inversely correlated with the DFS, indicating that they rapidly decay over time (Figure 2A). Follow up samples were collected from six recovered patients approximately 2-3 months following the previous blood draw time point and revealed a significant drop in anti-RBD levels reaching to the basal levels found in the control group (Figure 2B).

**Figure 2:**
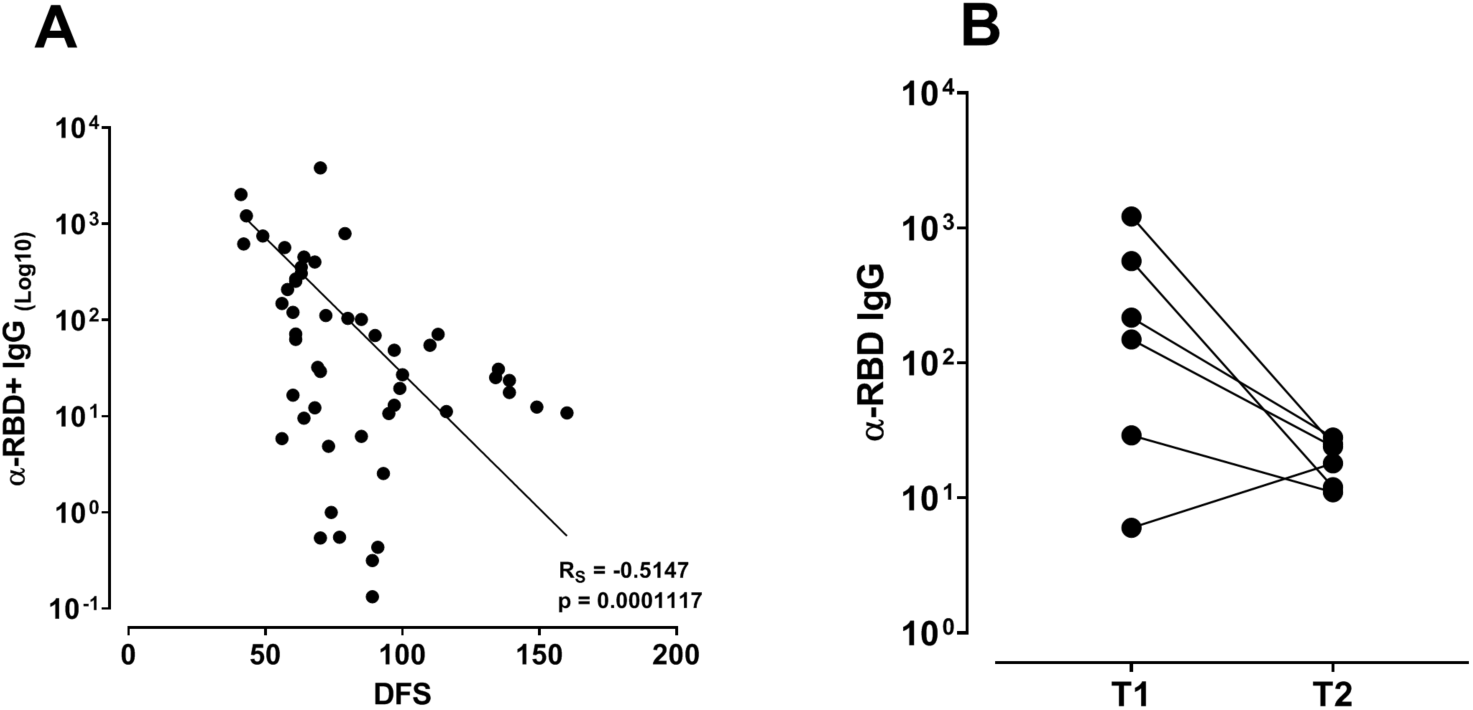
RBD-specific IgG dynamics in COVID-19 recovered patients. (A) Inverse correlation was found between RBD-specific IgG levels (n = 51) and DFS. Correlation was calculated using spearman’s rank correlation (R_S_). (**B**) The differential RBD-specific IgG levels were measured in follow up serum samples from Six recovered patients.

The “protective” capacity of the humoral memory arm following SARS-CoV-2 can be defined by the levels of antibodies specific for the pathogen and their neutralization ability. While, humoral memory, i.e., secretion of specific protective antibodies over time provides the host with a first line of defense against reinfection and is an informative marker for previous exposure. Because the half-life of secreted antibodies is short^24^, their persistence over time requires the presence of antibody secreting B cells (i.e. plasmablasts, plasma cells).

To monitor the B cell response in COVID-19 recovered patients and the association between RBD-specific B cell frequency with IgG levels, we applied PBMCs from recovered patients to FACS and isolated RBD-specific memory B cells (mBC) and plasmablasts (PB) using the double stain strategy (Figure S6).

To this end, we calculated the frequency of RBD-specific mBC and PB out of the total CD19^+^ cell population and analyzed the correlation between the two B cell subsets and with DFS and RBD-specific antibody levels. In contrast to the decay identified for the serological memory as reflected by the RBD-specific IgG, we observed that RBD-specific mBC were stable over time (Figure 3A). Follow up samples from a subset of the COVID-19 recovered patient cohort, demonstrated similar persistence of RBD-specific mBC and in some cases showing a mild increase of the RBD-specific mBCs frequency over time (Figure 3B). These data suggest that the B cell memory compartment is stable for at least 6 months following the recovery from COVID-19. These results are in accordance with the general dynamics observed in the mBC compartment where once established, antigen-specific memory B cell populations are remarkably stable and highly enriched for quiescent and exceptionally long-lived cells antigen-specific mBC^25^.

**Figure 3:**
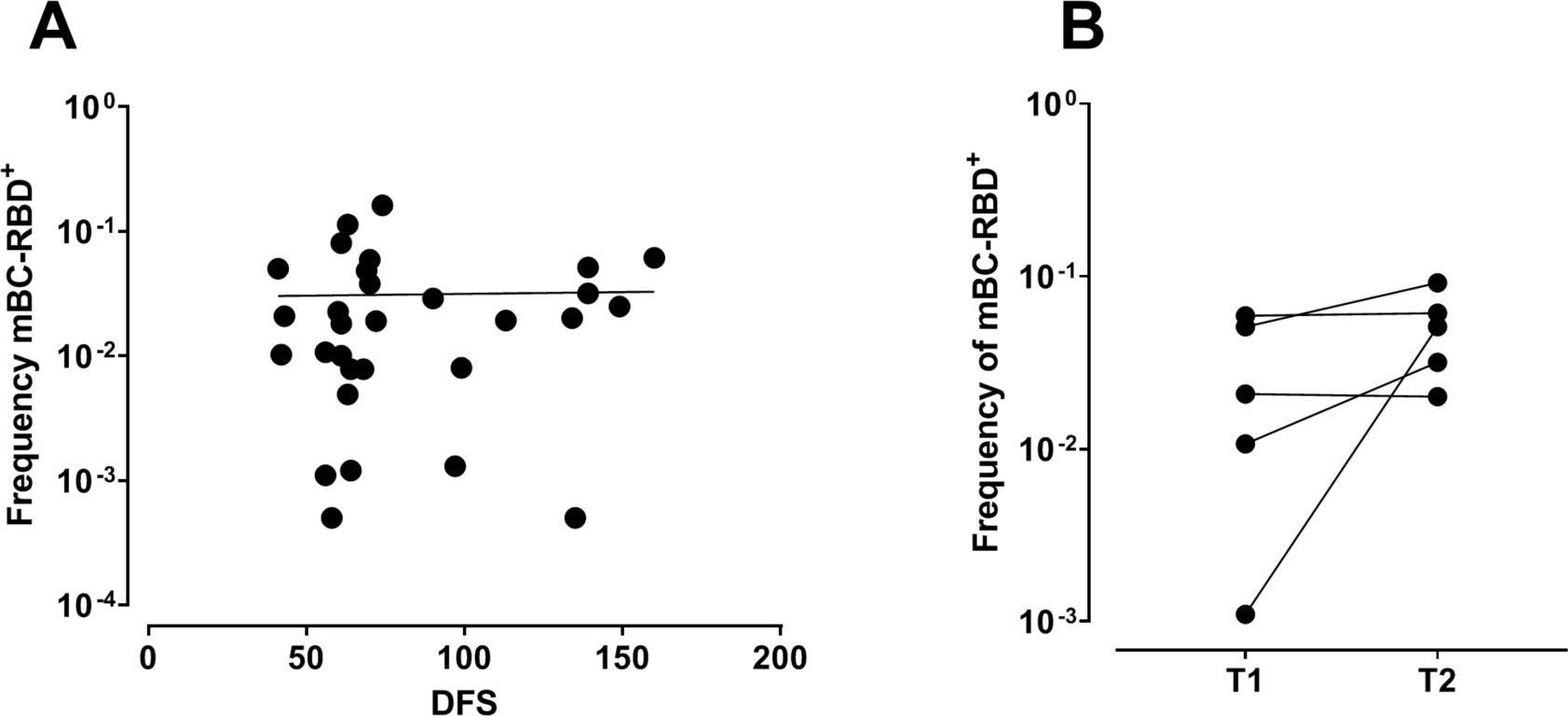
Dynamics of RBD-specific mBC. (A) No linear regression was identified for RBD-specific mBC over time (n = 30, R^2^ = 0.0004). (B) RBD-specific mBC frequencies in follow-up samples (n = 5) at two time points for COVID-19 recovered patients demonstrating that mBC within patients are stable over time.

It was recently reported that the B cell response following influenza vaccine does not induce long-lived plasma cells (LLPC) or these LLPC decline over time. Moreover, the associated influenza-specific antibodies decline in accordance with the LLPC frequency^26^. We hypothesized that the differentiation of B cells to become LLPC is impaired following SARSCoV-2 infection as we observed that antigen-specific antibodies decline over time. To test this hypothesis, we studied the association between the frequency of RBD-specific PB subset and the frequency of RBD-specific mBC and found that the frequency of both subsets are highly correlated (Figure 4A). These data are consistent with the immunological mechanism governing the B cell response following affinity maturation including asymmetrical deviation of parent B cell post germinal center affinity maturation^27^. Next, we investigated the association between RBD-specific PB and the antibody levels as such association will suggest that the main contributor for the serum antibody levels are the short lived plasmablasts (SLPB). Using linear regression analysis, we found an association between the RBD-specific IgG and the frequency of PB, suggesting that the contributor for serum IgG levels are mainly based on the presence of activated PB in the blood. In case LLPC were generated we would expect to find a decrease in PB frequency while a stable level of RBD-specific IgG.

**Figure 4:**
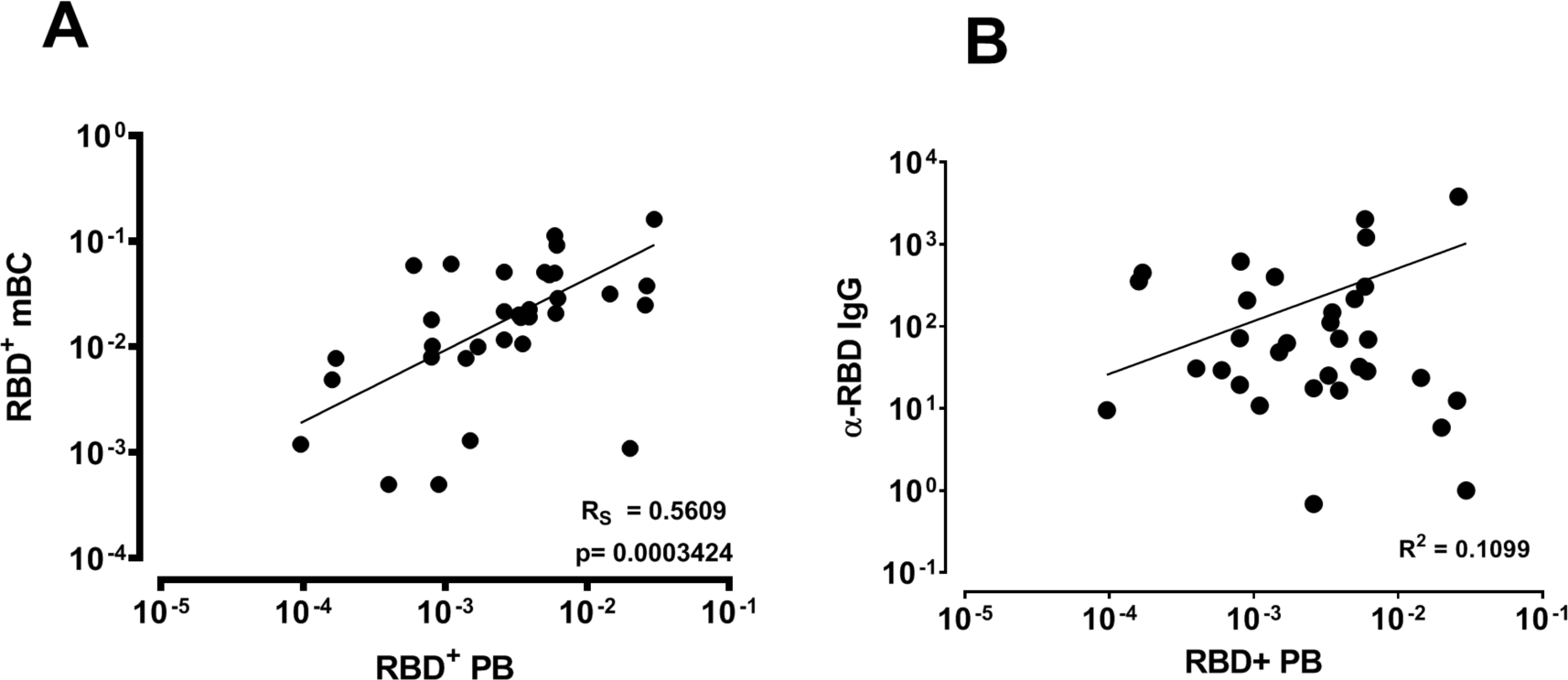
Correlation between RBD-specific PB and (**A**) RBD-specific mBC (n = 33) and (**B**) RBD-specific IgG (n = 31). For (A) spearman’s rank correlation was employed and for (B) linear regression, where R squared is designated.

The persistence of RBD-specific mBC suggests that the cellular arm of the adaptive immunity may provide robust recall immune response in case of re-infection however, the question remains - will this suffice in providing sufficient protection especially due to the decay of the serological memory? These aspects should be further investigated in follow up temporal studies when samples from recovered patients one year following onset of symptoms will be available.

### Materials and methods

#### Expression and purification of recombinant protein

The plasmids for expression of recombinant SARS-CoV-2 receptor-binding domain (RBD) was kindly provided by Dr. Florian Krammer, Department of Microbiology, Icahn School of Medicine at Mount Sinai, New York, NY, USA. The RBD sequence is based on the genomic sequence of the first virus isolate, Wuhan-Hu-1, which was released on January 10th 2020^5^. The plasmids for expression of recombinant human ACE2 (hACE2) was kindly provided by Dr. Ronit Rosenfeld from the Israel Institute for Biological Research (IIBR). The cloned region encodes amino acids 1-740 of hACE2 followed by a 8xHis tag and a Strep Tag at the 3’ end, cloned in a pCDNA3.1 backbone. Recombinant proteins were produced in Expi293F cells (Thermo Fisher Scientific) by transfections of these cells with purified DNA using an ExpiFectamine 293 Transfection Kit (Thermo Fisher Scientific), according to the manufacturer’s protocol, and as described previously^5^. Supernatants from transfected cells were harvested on day 6 post-transfection by centrifugation of the culture at 4000xg for 20 minutes and applied to a HisTrap affinity column (GE Healthcare) that was pre-equilibrated with binding buffer (PBS, pH 7.4, 5mM imidazole).

Affinity column was washed with 5 column volumes (CV) of wash buffer (PBS, pH 7.4, 20mM imidazole) followed by 2-step elution with 5CV of elution buffer (PBS, pH 7.4, 50 or 250mM imidazole). Elution was collected in 1ml fractions and were analyzed by 12% SDS–PAGE. Fractions containing clean recombinant proteins were merged and dialyzed using Amicon Ultra (Mercury) cutoff 10K against PBS (pH 7.4). Dialysis products were analyzed by 12% SDS– PAGE for purity and concentration was measured using Take-5 (BioTek Instruments).

Purified recombinant proteins were biotinylated using the EZ-Link Micro-NHS-PEG4-Biotinylation kit (Thermo Scientific), according to the manufacturer’s protocol. Biotinylated recombinant RBD was bound to Brilliant Violet 421^TM^ Streptavidin (BioLegend) or APC Streptavidin (Southern Biotech), as previously described^28^.

#### Blood samples

All patients provided informed consent to the use of their data and clinical samples for the purposes of the present study and blood collection was performed under institutional review board approvals number 0001281-4 and 0000406-1. All blood samples were collected at the Hasharon Hospital, Rabin Medical Center under ethical approval number 0265-20. Fifty-four blood samples from 42 active COVID-19 patients were collected into BD vacutainer serum collection tubes. Isolation of serum was performed according to the manufacturer’s protocol. Heat-inactivation of sera was performed by incubating the samples at 56°C for 30 minutes.

Fifty-seven blood samples from recovered patients and additional 6 follow up blood samples were collected into BD vacutainer K2-EDTA collection tubes. Isolation of serum and peripheral blood mononuclear cells (PBMCs) was performed by density gradient centrifugation, using Uni-SepMAXI+ lymphocyte separation tubes (Novamed) according to the manufacturer’s protocol. Control serum samples were from 26 healthy individuals. All serum samples were aliquoted and stored at −80°C.

#### Serum titer and serum neutralization capacity

Serum IgG, IgA and IgM antibody levels were measured by ELISA, as previously described^12,29^ with several modifications. Briefly, RBD-specific Ig levels in active patient sera were determined using 96 well ELISA plates that were coated overnight at 4°C with 2μg/ml RBD in PBS (pH 7.4). Next, coating solution was discarded and ELISA plates were blocked with 300µl of 3% w/v skim milk in PBS for 1 hour at 37°C. Following discarding blocking solution, duplicates of serum diluted 1:300 in 3% w/v skim milk in PBS were added to the wells. Negative control serum samples were also added in duplicates of 1:300 dilution. ELISA plates were washed three times with PBST and 50μl of horseradish peroxidase (HRP) conjugated anti-human IgG H+L / anti-human IgM / anti-human IgA secondary antibodies were added to each plate at the detection phase (50μl, 1:5000 ratio in 3% w/v skim milk in PBS) and incubated for 1 hour at room temperature (RT), followed by three washing cycles with 0.05% PBST. Developing was carried out by adding 50µl of TMB and reaction was quenched by adding 0.1M sulfuric acid. Plates were read using the Epoch Microplate Spectrophotometer ELISA plate reader using wave length of 450 and 620 nm. The neutralization capacity of serum from recovered patients was determined using 96 well ELISA plates that were coated overnight at 4°C with 2μg/ml RBD in PBS (pH 7.4). Next, coating solution was discarded and ELISA plates were blocked with 300µl of 3% w/v skim milk in PBS for 1 hour at 37°C. Blocking solution was discarded and 50µl of 400nM hACE2 in 3% w/v skim milk were added to the positive hACE2 wells, and 3% w/v skim milk in PBS was added to the negative hACE2 wells for 1 hour at RT. Next, triplicates of 1:150 diluted serum samples w/ and w/o 400 nM hACE2 were added to the positive/negative hACE2 wells (respectively) and serially diluted 3-fold in 3% w/v skim milk in PBS (1:150– 1:328,05 serum dilution factor). Negative serum samples were also added in duplicates of 1:150 dilution w/ and w/o 400nM hACE2. Plates were incubated for 1 hour at RT. ELISA plates were washed three times with PBST and 50μl of HRP conjugated anti-human IgG H+L secondary antibody were added at the detection phase (50μl, 1:5000 ratio in 3% w/v skim milk in PBS) and incubated for 1 hour at RT, followed by three washing cycles with 0.05% PBST. Developing was carried out by adding 50µl of TMB and reaction was quenched by adding 0.1M sulfuric acid. Plates were read using the Epoch Microplate Spectrophotometer ELISA plate reader using wave length of 450 and 620nm.

#### Flow cytometry analysis

PBMCs from COVID-19 recovered patients were stained for 15 minutes in cell staining buffer (BioLegend) at RT in the dark using the following antibodies: anti-CD19–APC/Cyanine7 (clone HIB19; BioLegend), anti-CD20–FITC (clone 2H7; BioLegend), anti-CD27–PerCP/Cyanine5.5 (clone O323; BioLegend), anti-CD38–PE (clone HB7; BioLegend), biotinylated recombinant RBD coupled to Brilliant Violet 421^TM^ Streptavidin (RBD-BV421), and biotinylated recombinant RBD coupled to APC Streptavidin (RBD-APC).

The following B cell population was sorted using a BD Aria III cell sorter (BD Bioscience): PB (CD19^+^CD20^−^CD27^high^CD38^high^), RBD specific PB (CD19^+^CD20-CD27^+^CD38^high^RBD^+(double stained)^), mBC (CD19^+^CD20^+^CD27^var^) and RBD specific mBC (CD19^+^CD20^+^CD27^var^RBD^+(double stained)^). Gating strategy used for cell sorting was consistent throughout all samples, on singlets that were CD19+. B cells frequencies were calculated using FlowJo v10 software (BD, USA).

### Statistical analysis

All curves were fitted on a sigmoidal dose–response curve and area under curve of each was calculated. Mann-Whitney test was used to compare continuous variables. All reported P values were two-tailed, and a P value less than 0.05 were considered statistically significant. All statistics were performed with GraphPad Prism software (version 8, San Diego, California).

## Data Availability

The data that support the findings of this study are available from the corresponding author, [YW], upon reasonable request.

## Acknowledgment

We wish to thank Prof. Itai Benhar and Dr. Limor Nahary from the Shmunis school of Biomedicine and cancer research, Tel Aviv University, for their assistance in providing the initial RBD and hACE2 reagents, Dr. Ronit Rosenfeld from the Israel Institute for Biological Research (IIBR) for her generous assistance and Prof. Mordechai Gerlic and Prof. Ariel Munitz from the Sackler Faculty of medical, Tel Aviv University.

---

**Figure S1:**
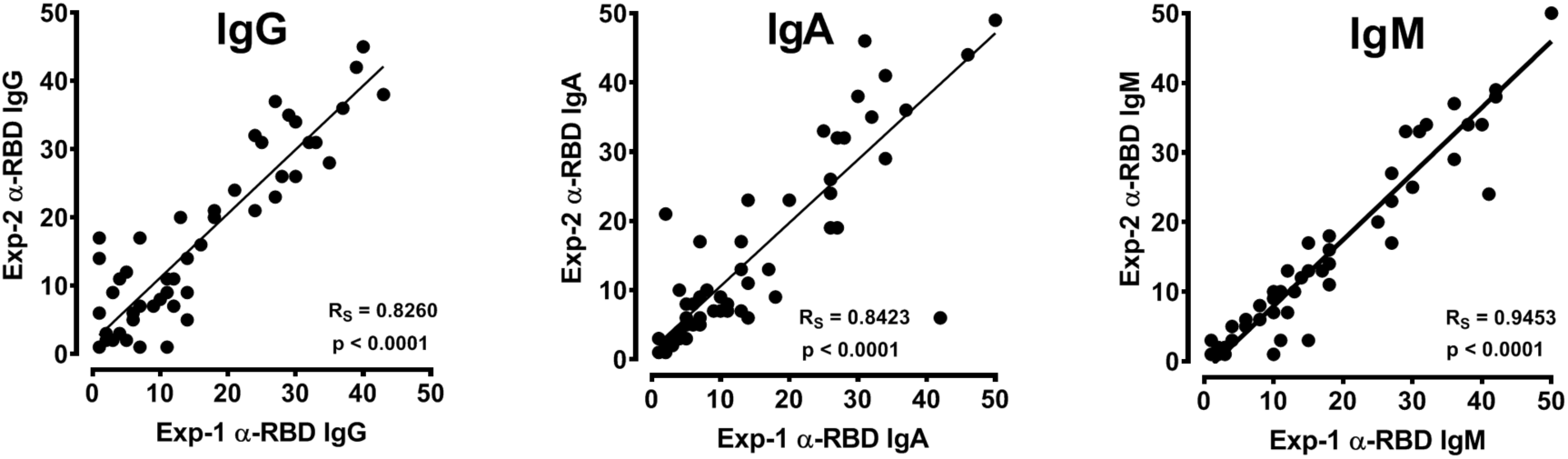
Reproducibility of the ELISA experiments to determine RBD-specific antibodies in symptomatic COVID-19 patients. Each experiment included negative controls and antibody levels were calculated as the signal over background ratio. The values were ranked (x-axis) according to the antibody relative levels and applied to spearman’s rank correlation. Rank correlation is designated (R_S_) and p values for each antibody isotype.

**Figure S2:**
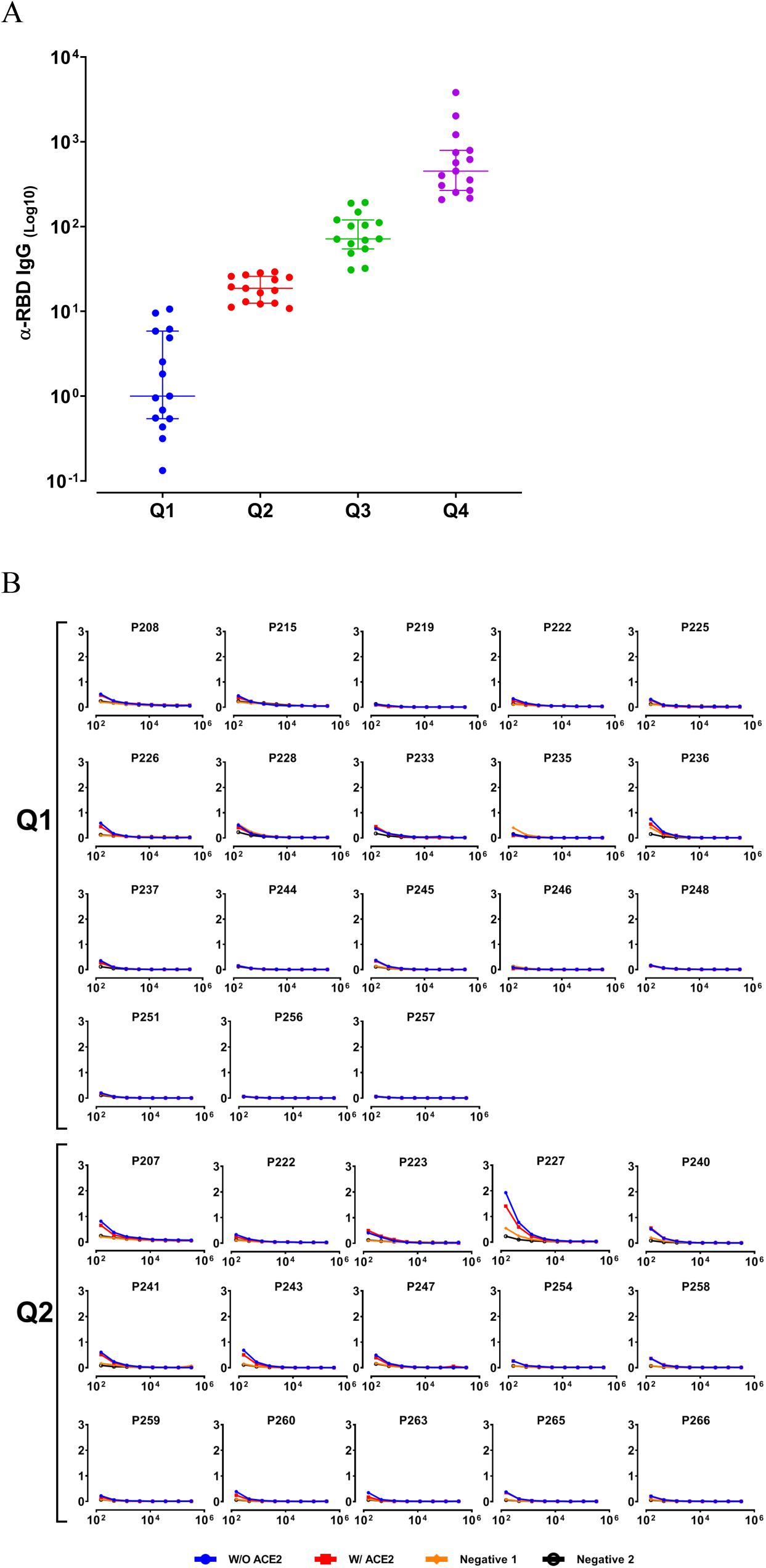
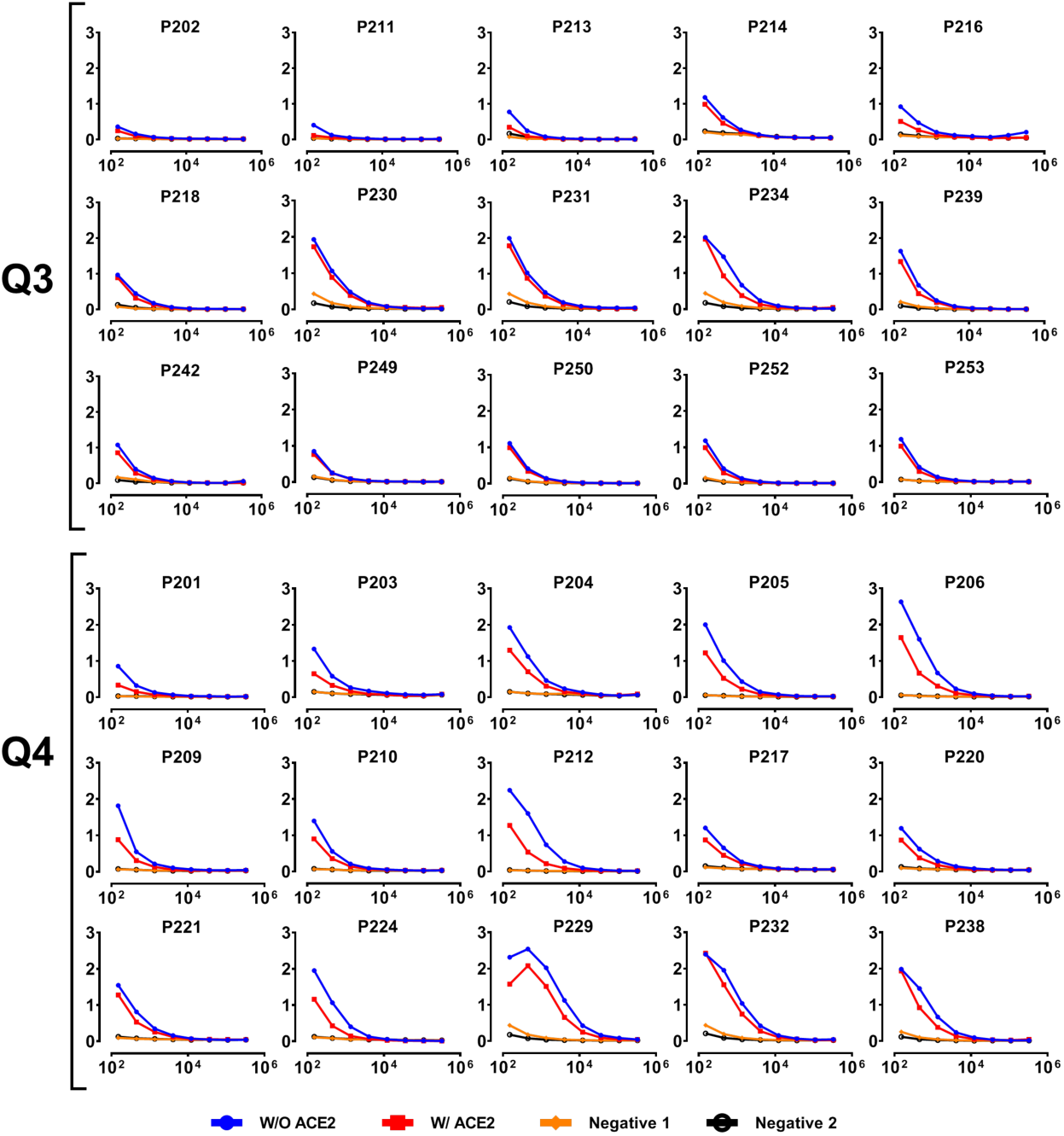
Total RBD-specific IgG and neutralizing IgG in COVID-19 recovered patients as determined by in vitro neutralization assay. For each serum, the differential area under curve (AUC) was calculated for the total RBD-specific IgG and neutralizing IgG. The ratio between the mean AUC of the control sera and the tested serum was calculated and stratified to quartiles (A). Specific serum dilution ELISA for all sample (B) w/ and w/o hACE2 as the competitive reagent.

**Figure S3:**
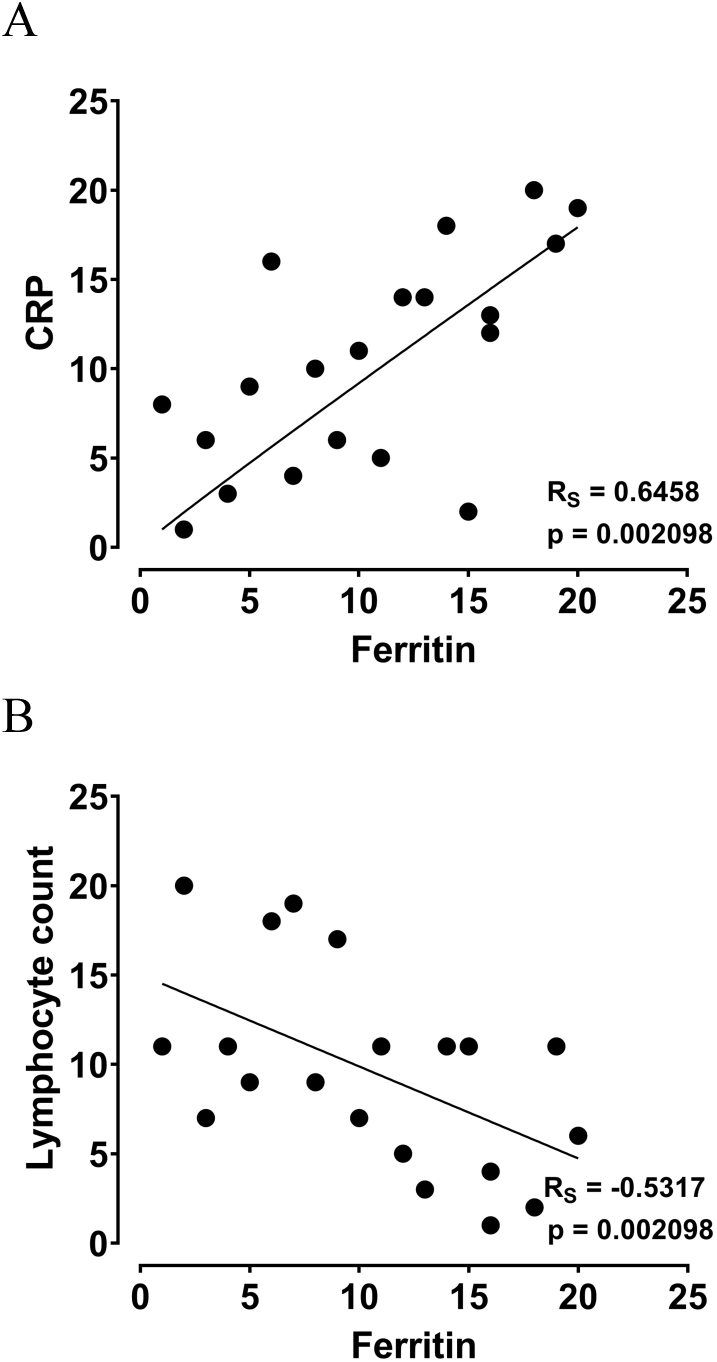
Correlation between inflammatory risk factors CRP, Ferritin and lymphocytes counts. All values were normalized to ranking values correlation was calculated using spearman’s rank correlation based on the original values. Lymphocytes – 1K/µl, CRP – mg/dl and ferritin – µg/ml. (**A**) CRP was highly correlated with ferritin levels in blood while (**B**)lymphocytes were inversely correlated with ferritin levels. R_S_ – spearman’s rank correlation. Linear regression line is presented as a solid line.

**Figure S4:**
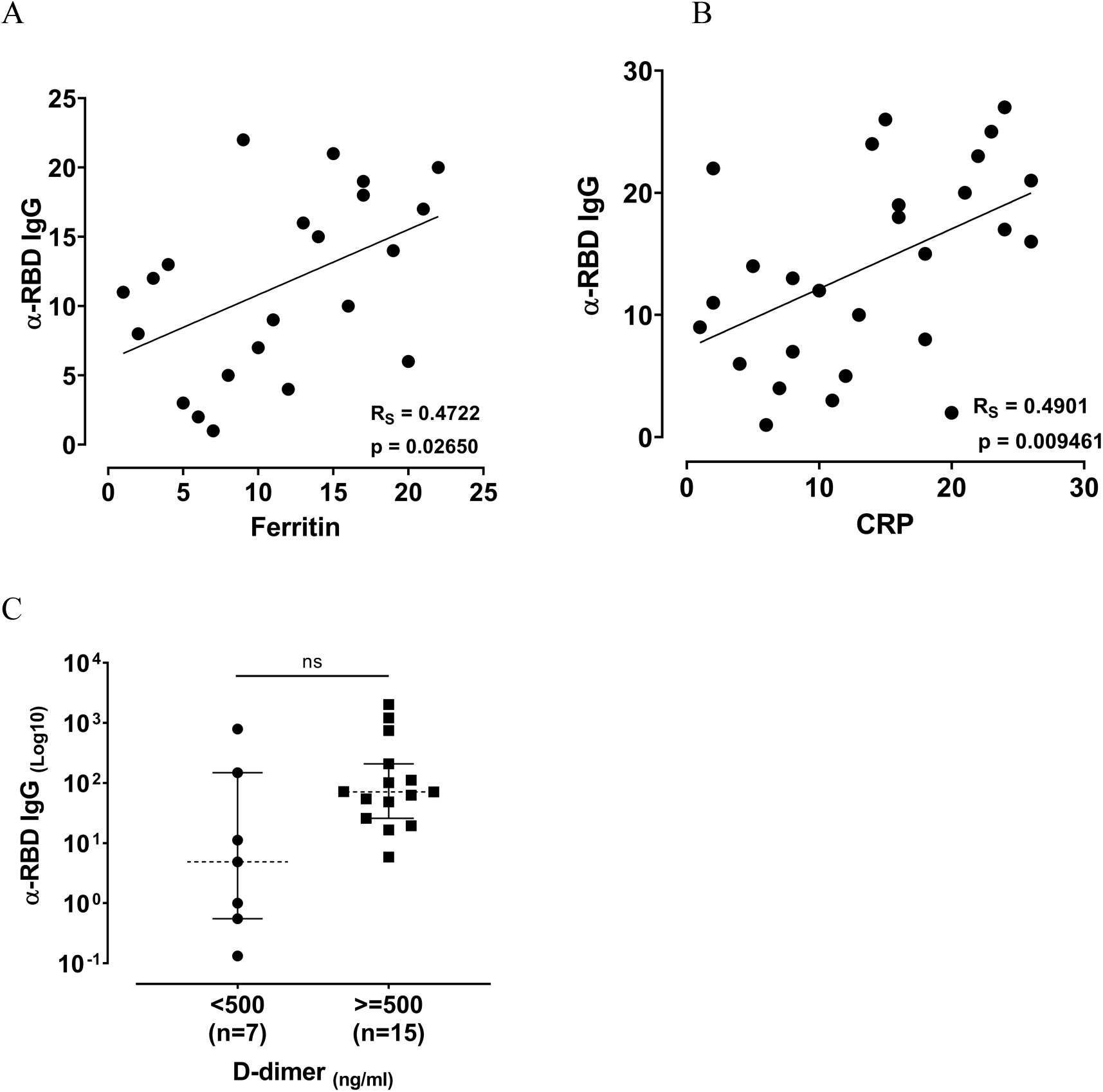
Correlation between inflammatory risk factors CRP, Ferritin. All values were normalized to ranking values correlation was calculated using spearman’s rank correlation based on the original values. CRP – mg/dL and ferritin – µg/ml. CRP (**A**) and ferritin (**B**) were found to be correlated with RBD-specific antibody levels. R_S_ – spearman’s rank correlation. Linear regression line is presented as a solid line. The association between D-dimer and RBD-specific antibodies was tested using nonparametic Mann-Whitney test. (ns p > 0.05).

**Figure S5:**
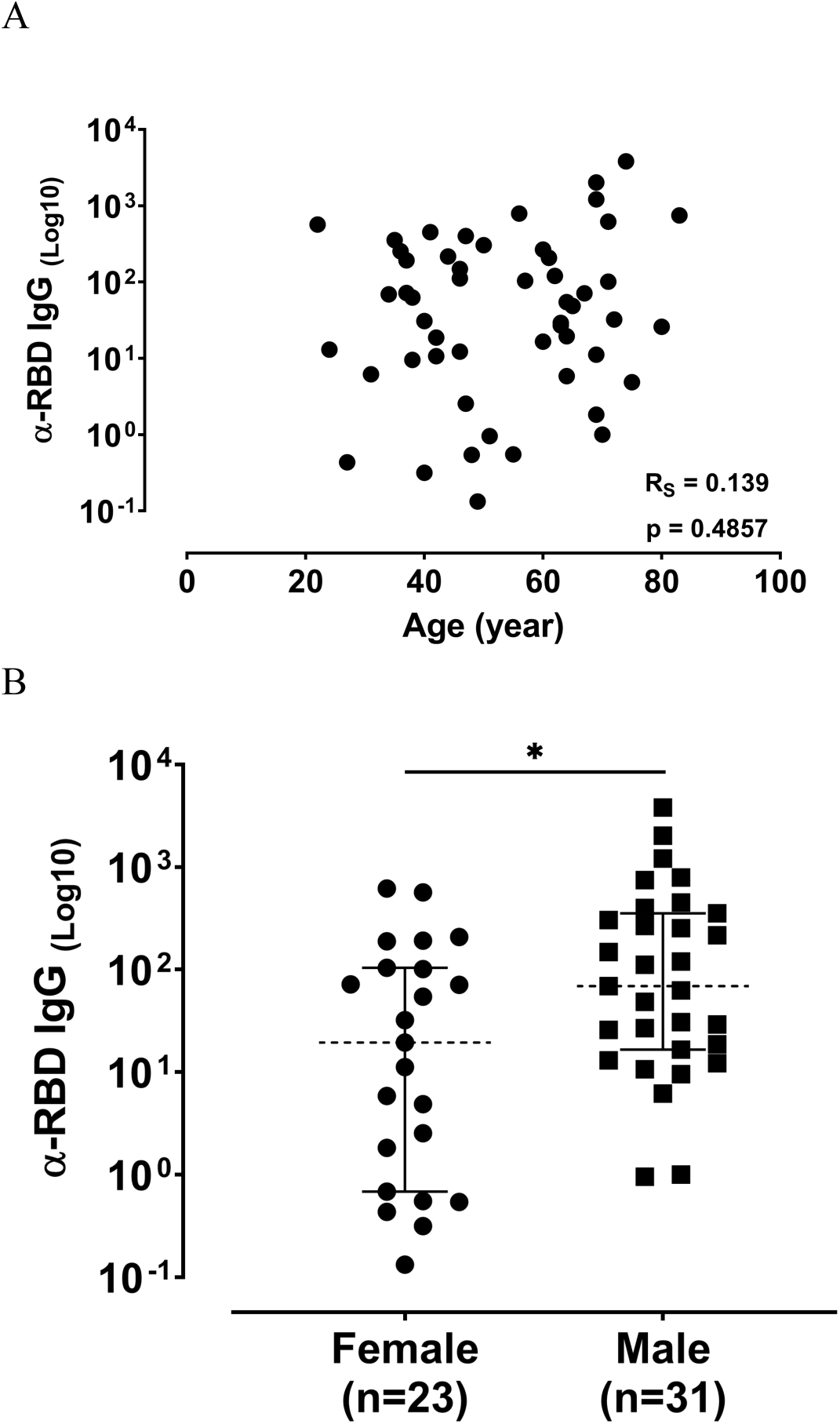
Association between age (**A**), gender (**B**) and RBD-specific antibodies in recovered COVID-19 patients. Age association was tested by spearman’s rank correlation (R_S_) and gender by nonparametic Mann-Whitney test. (ns p > 0.05, * p ≤ 0.05).

**Figure S6:**
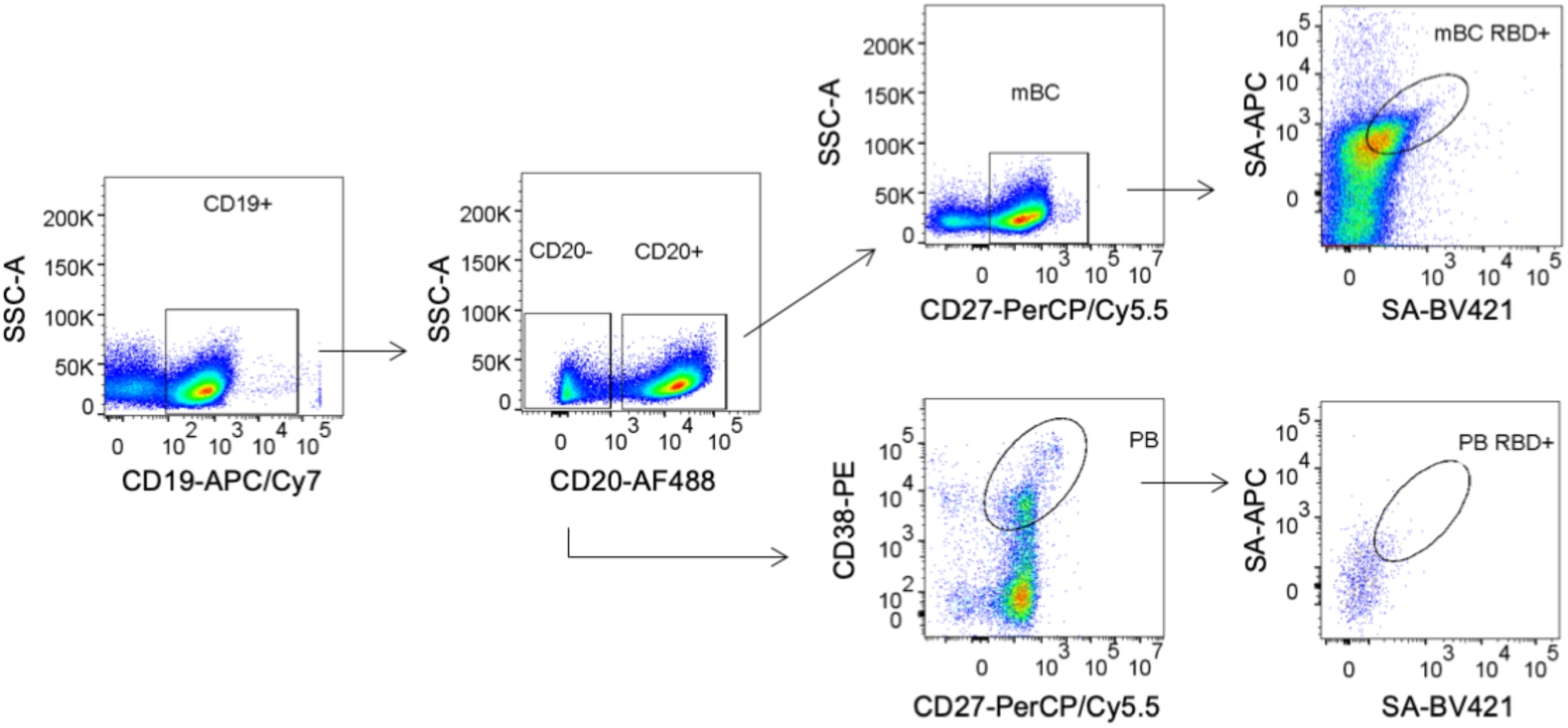
Representative example of fluorescence-activated cell sorting (FACS) of PBMC from COVID-19 recovered patient. PBMCs were enriched for B cells by negative selection using EasySep magnetic beads followed by FACS to isolate antigen-specific mBC (upper panel - CD19^+^CD20^+^CD27^var^RBD^double positive^) and antigen-specific plasmablasts (lower panel – CD19^+^CD20^−^CD27^high^CD38^high^RBD^double positive^). Frequencies of B cell subsets were calculated per total CD19^+^ B cells.

**Table S1:** Summary of enrolled patients to the study. For each parameter the number of patients is different as not all data was available. Over all 54 symptomatic patients and 57 recovered patients were enrolled. 26 control sera sample were used from a retrospective serum bank available in the laboratory.

|  | Symptomatic | Recovered |
| --- | --- | --- |
| Median ages | 69 (35-95), n=54 | 55.5 (22-83), n=56 |
| Gender |  |  |
| Female | 32.07% (n=17) | 48.3% (n=29) |
| Male | 67.93% (n=36) | 51.7% (n=31) |
| Median DFS | 13 (0-60), n=53 | 70 (41-135), n=45 |
| Laboratory findings |  |  |
| CRP (mg/dL) | 2 (0-34.5), n=51 | 1.2 (0.1-36), n=27 |
| Lymphocytes (1000/μL) | 1.2 (0.5-10.5), n=51 | 1.1 (0.4-3.3), n=35 |
| Ferritin (μg/mL) | NA | 508.5 (11-2865), n=22 |
| D-Dimer (ng/mL) | NA | 836 (109-8028), n=21 |
| Underlying medical conditions |  |  |
| Hypertension | NA | 39.6% (n=48) |
| Dyslipidemia | NA | 41.7% (n=48) |

